# Surveillance and Stability of SARS-CoV-2 Wastewater Samples in Minnesota

**DOI:** 10.1101/2023.10.14.23296666

**Authors:** Mark J. Osborn, Shannon Champeau, Carolyn Meyer, Mason Hayden, Laura Landini, Stacey Stark, Stephanie Preeket, Sara Vetter, Ruth Lynfield, Daniel Huff, Timothy W. Schacker, Charles R. Doss

**Affiliations:** Department of Pediatrics, University of Minnesota, Minneapolis, MN; U-Spatial, University of Minnesota-Duluth, Duluth, MN; Minnesota Department of Health, St Paul, MN; Department of Medicine, University of Minnesota, Minneapolis, MN; School of Statistics, University of Minnesota, Minneapolis, MN

## Abstract

Wastewater-based epidemiology provides an approach for assessing the prevalence of pathogens such as COVID-19 in a sewer service area. In this study, SARS-CoV-2 RNA was measured serially in 44 wastewater treatment plants of varying service capacities comprising approximately 67% of the population of Minnesota, from September 2020 through December 2022. We employed linear regression models to establish a predictive relationship between the weekly SARS-CoV-2 RNA concentrations in wastewater and clinical case counts. Metrics were assessed under specified transformation and normalization methods which we confirmed by cross-validation averaged across the enrolled treatment plants. We report that the relationship between COVID-19 incidence and SARS-CoV-2 RNA in wastewater may be treatment plant-specific. Toward establishing guidelines for pathogen surveillance, we further studied storage and time-to-analysis for RNA wastewater data and observed large effects of storage temperature, indicating that collection methods may have an important effect on the utility and validity of wastewater data for infectious disease monitoring. Our findings are additive for any large-scale wastewater surveillance program.

## 1 Introduction

Coronavirus disease 2019 (COVID-19), caused by severe acute respiratory syndrome coronavirus 2 (SARS-CoV-2), has infected millions of people across the globe which continues to cause significant health and economic impacts, and demonstrated the importance of having infrastructure available to deploy for monitoring diseases on a massive scale. Effective and widespread community testing of individuals is costly and the demand for tests frequently exceeds the capacity of testing facilities (Barasa, Ouma, and Okiro 2020), and there is a disequilibrium in access to testing due to economic, geographic, or social conditions. Furthermore, test results are a lagging indicator of the pandemic’s progression, because testing is usually prompted by symptoms, which may take days or weeks to manifest after infection (Lauer et al. 2020). Furthermore, with broader use of home testing, the less reliable official clinical testing becomes, since those not requiring medical attention will tend to also avoid medical facilities that report public health testing data. Thus, delays may occur between the appearance of symptoms, testing and the reporting of test results (Peccia et al. 2020). Finally, it is estimated that as many as 45% of COVID-19 cases are asymptomatic (Li et al. 2020; Nishiura et al. 2020; Oran and Topol 2020; Post et al. 2020). Considering that most people only seek medical attention and undergo diagnostic testing if they are symptomatic, the number of confirmed clinical cases may grossly underestimate the prevalence of the disease in a community.

Wastewater-based epidemiology (WWBE) is a method for monitoring presence and trends of an infectious agent that is shed in bodily secretions/excrement in communities. In WWBE for SARS-CoV2, wastewater is collected from wastewater treatment plants (WWTPs) and is tested by qRT-PCR for SARS-CoV-2 RNA excreted via feces, urine, and saliva. The presence and levels of viruses in wastewater samples represents a cross section of viral presence and spread in the communities served by those plants. WWBE has been successfully employed as a surveillance tool for diseases such as SARS, hepatitis A, and polio (Hellmér et al. 2014; Manor et al. 1999; Ye et al. 2016). With regard to SARS-CoV-2, viral particles are reported to be shed in feces from infected individuals even if they are asymptomatic (Chan et al. 2021; Chen et al. 2020; Cheung et al. 2020; Parasa et al. 2020; Wong et al. 2020). Recent studies have shown that WWBE is able to predict COVID-19 prevalence even earlier than clinical case data Peccia et al. (2020), supporting the idea that WWBE can be used as an early warning system to monitor community prevalence and spread.

Over the last three years we have gathered SARS-CoV-2 wastewater data, which we present in this paper both to describe the programmatic data to date and to provide more about the specifics of the SARS-CoV-2 pandemic, and to identify best practices for WWBE more broadly. We present data from two complementary data gathering processes here. The primary data is wastewater samples that were collected from 44 total WWTPs across the state of Minnesota between September 2020 and December 2022. The SARS-CoV-2 viral load in wastewater were obtained by qRT-PCR. These 44 WWTPs represent a broad sampling of the Minnesota population serving a total of 191 zip codes and a population of 3,825,269 people, which is approximately 67% of the total population of the state. The secondary data were from an experiment on storage and time-to-analysis conditions for qRT-PCR analysis of wastewater data. This experiment was undertaken in part because of our findings from our primary data of significant heterogeneity in the data quality and predictive ability from different WWTPs.

Estimating the SARS-CoV-2 RNA concentrations in wastewater (gene copies per litre) is complicated, as the dilution and fecal strength in the wastewater may vary between sampling dates due to random chance (e.g., caused by variation in a factory’s runoff). It has been recommended to multiply the viral concentration in wastewater by the flow of the sampled location (the volume of wastewater that passed through the location in a day) to obtain the viral concentrations in gene copies per day, and account for changes in sanitary sewer contributions (Hasan et al. 2021; Weidhaas et al. 2021). Normalizing SARS-CoV-2 RNA concentrations by indicators of human fecal waste is also common, because feces in wastewater can have variable levels of SARS-CoV-2 depending upon the amount of water used per toilet flush or body washing (Zhan et al. 2022). The contribution of SARS-CoV-2 from human sourced water can then be estimated by dividing the measured SARS-CoV-2 concentration by the concentration of the human waste indicator (Zhan et al. 2022). One typically examined fecal marker is Pepper Mild Mottle Virus (PMMoV) Maal-Bared et al. (2023). Previous studies have shown that PMMoV is the most abundant RNA virus in human feces and it is shed in large quantities in wastewater (Melvin et al. 2021, Hamza et al. 2019; Kitajima et al. 2014; Kitajima, Sassi, and Torrey 2018; Rosario et al. 2009; Zhang et al. 2006). It is also highly stable in wastewater, and its concentrations show little seasonal variation (Kitajima et al. 2014; Kitajima, Sassi, and Torrey 2018). Operating on this information, our program from 2019-2022 normalized SARS-CoV-2 RNA wastewater levels to PMMoV (Melvin et al. 2021). However, in our ongoing surveillance work, we found that normalizing by PMMoV obfuscated the large spike in wastewater measurements during the COVID-19 Omicron wave of December 2021-January 2022, which compelled us to reexamine normalization methods. Our findings below suggest that normalization by PMMoV may increase the variation in wastewater measurements, and we do not find evidence in the current data that it substantially improves prediction.

Our study objectives in conducting these analyses were to: (1) learn how best to make use of large-scale wastewater surveillance RNA measurements, and (2) storage conditions that affect said measurements. Our motivation for (2) was driven in part by our results in (1), in which we found nontrivial unexplained heterogeneity between WWTPs. We compare a variety of data pre-processing methods; to benchmark the effectiveness of a pre-processing method we use predictive ability of the data on clinical case counts. We assess model fit across a variety of models (and data transformations) by using cross-validation (see the methods section).

However, we have observed nontrivial heterogeneity in the predictive ability of measurements from differing WWTPs. Researchers have started to investigate how storage conditions affect wastewater RNA and measured concentrations (Khan, Tighe, and Badireddy 2021). In a small scale study focusing on one or a small number of WWTPs, the storage and measurement conditions can easily be controlled. As we will discuss in our results section, we observed significant heterogeneity in the relationship of the wastewater measurements with clinical COVID-19 case counts which we attribute to heterogeneous quality of data from the WWTPs. Thus, we also implemented a study on storage and time-to-analysis conditions for measurements from wastewater, to undertake efforts how much these aspects affect quality of measurements in order to define sample collection guiding principles.

In addition to this study on wastewater measurement quality, we study the following questions about statistical processing and analysis of wastewater data. Rather than use statistical significance testing of model coefficients, or coefficient of determination (*R*^2^) from linear regression for assessment of fit, we will use *leave-one-out cross-validation*, which is appropriate for assessing model performance when prediction is the main goal. (See the methods section for a full description.)

## 2 Description of Surveillance Data

Wastewater samples were collected from a total of 44 WWTPs across the state of Minnesota. From its inception to March 2022, we employed a polyethylene glycol concentration and phenol chloroform nucleic acid extraction procedure and qRT-PCR for the nucleocapsid gene in the SARS CoV2 genome using N1 and N2 primer:probe pairs. As the pandemic progressed so too did more sensitive and effective isolation and detection methods. Accordingly, in March 2022 we pivoted to a column-based total nucleic acid isolation method and a three gene qRT PCR method that measured the nucleocapsid (N), spike (S), and ORF1ab (O) proteins, sampled from 40 WWTPs. Our statistical findings demonstrate the latter assays are more accurate than the former assays. We present here, data from the latter period, using the three gene qRT PCR assay. Since the two assays are nontrivially different, they cannot be combined into a single statistical analysis. For data analysis purposes, a weekly level average of measurements was used. Figure 2.1 displays the concentrations of N, S and O in three WWTPs, Little Falls, Northfield and Twin Cities, which are chosen as representatives of smaller, midsized, and large population areas (see Table 2.1 below). One sample was reported to have zero concentration of N, S and O. It was removed in further analyses. Among the remaining samples, 424 were reported to have zero virus concentration of S. The reason for the loss of signal (S gene target failure) was due to prevalence of a SARS CoV2 variant that acquired mutations in the spike gene that prevented the primer:probe pair from binding. Thus, the S gene was not used in any further analyses. Importantly, our analytical resolution was preserved using the N and Orf1a genes. The concentrations of a human fecal marker, PMMoV, were also determined in each wastewater sample in parallel. PMMoV concentrations below the 5th percentile were replaced with the 5th percentile value, and those above the 95th percentile were replaced with the 95th percentile value to avoid outliers. The influent flow rate was provided by the participating WWTPs. The catchment area of each WWTP was obtained from the Minnesota Pollution Control Agency (MPCA) or, if possible, directly from the treatment facility. Many of the smallest WWTPs did not have a digitized catchment area available, so the city boundaries were used to approximate this area. Data enrichment services from Esri (ESRI 2023) were used to apportion 2020 census block population to the WWTP catchment areas.

**Figure 2.1:**
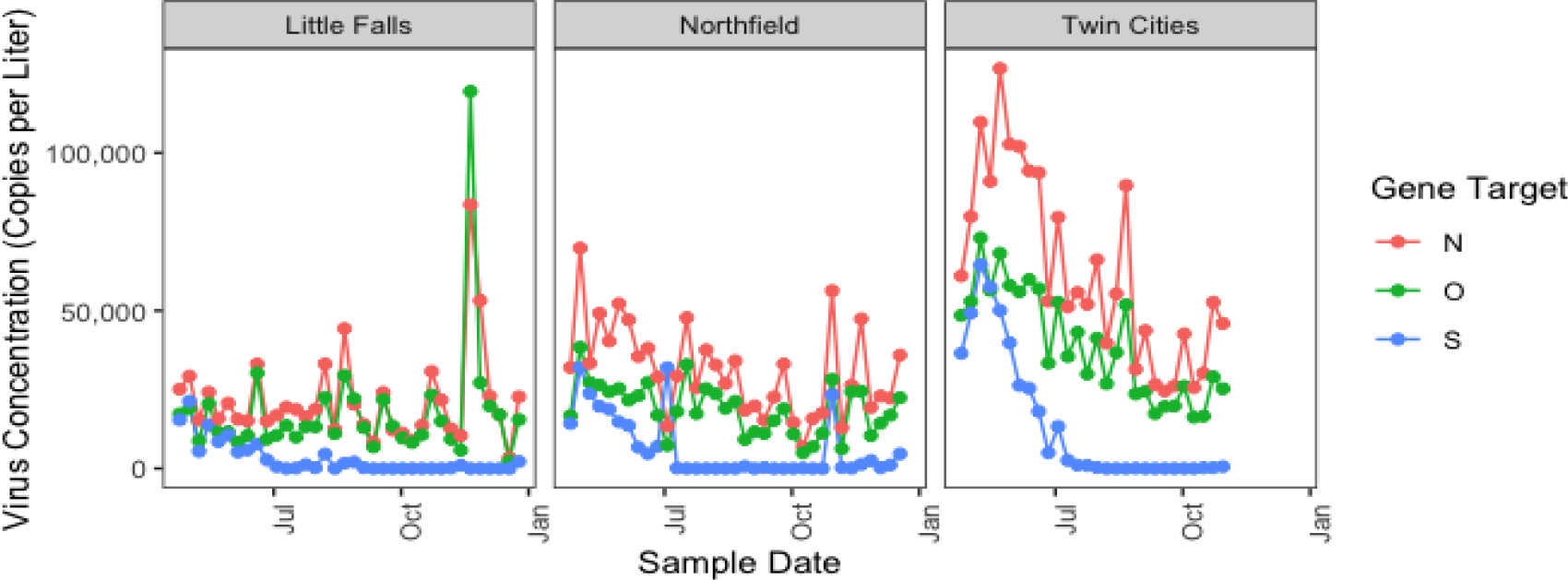
Virus concentrations in Little Falls, Northfield and Twin Cities

**Table 2.1:** Descriptions of participating WWTPs.

| **WWTP** | **Code** | **No.**<br>**Weeks**<br>**w/ data** | **Population** | **Avg. Weekly**<br>**Case Count** |
| --- | --- | --- | --- | --- |
| Albert Lea | AL | 36 | 20,957 | 28 |
| Alexandria | AX | 35 | 25,689 | 41 |
| Bemidji | BI | 26 | 32,741 | 38 |
| Blooming Prairie | BP | 10 | 3,589 | 4 |
| Blue Lake | BL | 28 | 397,299 | 674 |
| Cambridge | CB | 35 | 15,529 | 19 |
| Chisholm | CM | 20 | 5,787 | 9 |
| Eagles Point | EP | 29 | 113,786 | 208 |
| East Bethel | EB | 27 | 14,880 | 17 |
| Elk River | ER | 34 | 94,119 | 133 |
| Empire | EM | 28 | 176,441 | 287 |
| Fergus Falls | FF | 35 | 18,799 | 24 |
| Glacial Lakes SSWD | GL | 33 | 5,574 | 6 |
| Glencoe | GC | 9 | 8,280 | 14 |
| Hastings | HS | 28 | 30,027 | 39 |
| Hinckley | HY | 30 | 5,483 | 7 |
| Hutchinson | HT | 24 | 17,829 | 32 |
| International Falls | IF | 33 | 9,618 | 20 |
| Lafayette | LY | 15 | 844 | 1 |
| Lanesboro | LB | 24 | 1,849 | 2 |
| Le Sueur | LS | 35 | 8,237 | 7 |
| Little Falls | LF | 36 | 14,615 | 26 |
| Mankato | MK | 25 | 63,489 | 86 |
| Marshall | ML | 36 | 15,550 | 19 |
| Moorhead | MH | 31 | 42,602 | 69 |
| Mora | MR | 36 | 9,872 | 15 |
| New Prague | NP | 36 | 12,557 | 14 |
| North Branch | NB | 35 | 16,827 | 22 |
| Northfield | NF | 35 | 27,040 | 32 |
| Rochester | RC | 36 | 123,041 | 242 |
| Rogers | CR | 27 | 14,573 | 21 |
| Saint Cloud | SD | 35 | 130,901 | 260 |
| Seneca | SN | 28 | 243,338 | 430 |
| St. Croix Valley | SV | 28 | 38,958 | 68 |
| Thief River Falls | TR | 36 | 12,971 | 17 |
| Twin Cities | TC | 28 | 1,854,291 | 3,261 |
| Western Lake Superior | WL | 28 | 124,459 | 241 |
| Willmar | WM | 36 | 23,697 | 35 |
| Winona | WA | 34 | 34,644 | 59 |
| Worthington | WT | 36 | 14,487 | 18 |

**Figure 2.1:**
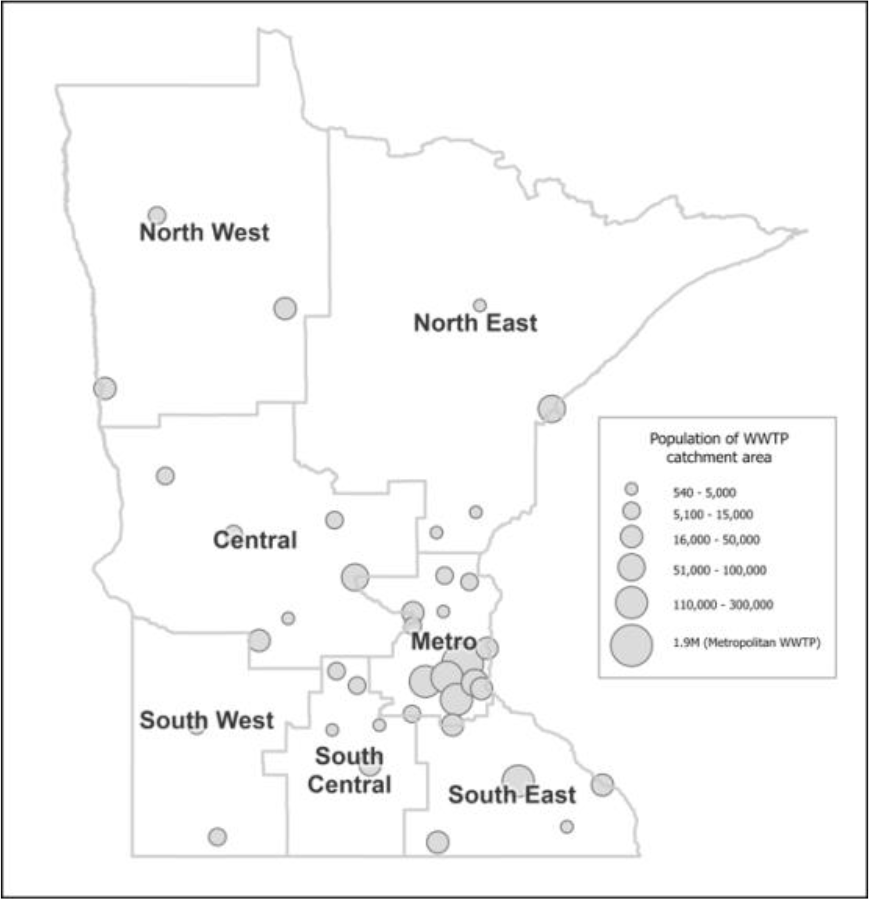
Map of WWTPs across the state of Minnesota.

The weekly number of new infections for each WWTP service area were obtained from the Minnesota Department of Health (MDH) and aggregated to the zip code areas using the residential address for the case. Case counts were summed by zip code weekly (Sunday-Saturday) and the date beginning the week was used to associate with each case count. The case data were then aggregated by WWTP according to the zip codes previously found to be associated with each catchment. Five zip codes intersected more than one WWTP catchment area. For the purpose of this study, the catchment area with more area in each zip code was assumed to serve the population (and therefore be the source of all the cases) of that entire zip code. To allow a log_10_ transformation, any zero case counts were replaced with a value of one. The MDH also provided the number of people who have received at least one dose of COVID-19 vaccine, and the number of people with completed vaccine series in the areas served by each WWTP over time. The complete series could be one, two, or three doses depending on the person’s age and which vaccine they received. Being “fully vaccinated” does not include or require further booster doses in the present definition.

Table 2.1 provides descriptions of the participating WWTPs, including the sampling period, the number of weeks where the WWTP was included in the study (because we had clinical case count data and had or could impute wastewater measurements), the size of the population served, as well as the sample means and standard deviations of weekly case count and flow.

## 3 Analysis of surveillance data

### Viral load quantification

The procedure we used for quantifying viral load is as follows: 40 mL of influent was used to isolate total nucleic acids using the Enviro Total Nucleic Acid Kit for Wastewater (Promega, Madison, WI) and eluted in a volume of 40 ul. Five ul was employed for quantitative RT PCR, performed in duplicate, using the TaqPath™ COVID-19 Combo Kit (ThermoFisher, Waltham, MA) with copy number standards on the QuanStudio 5 RT PCR instrument in order to quantitate viral copy number.

### Statistical Methods

The wastewater samples were collected at irregular time intervals and were isolated on different days for different WWTPs and COVID-19 case counts were aggregated weekly. To align the wastewater samples with the clinical case counts and to avoid having to analyze data on irregular time intervals, we aggregated the wastewater measurements by week (Sunday-Saturday). For a given week with any wastewater data, we use the sample average of all the measurements of that week as the aggregated measurement. If there was no wastewater sample in a week then that week was considered missing and was not included in the analysis.

With time-aligned case counts and wastewater measurements, we performed linear regression and fit separate models for each WWTP. To assess the model performance, a leave-one-out cross-validation (LOOCV) was used. The models were trained on *n* − 1 observations and validated on the remaining one observation, where *n* is the sample size. The procedure was repeated *n* times with each of the *n* observations used exactly once for validation. The average of the *n* prediction errors obtained was computed for model comparison. The evaluation metric was the root mean squared errors (RMSE) between the predicted value and the actual value. If the dependent variable was log_10_-transformed, the transformation was reversed to obtain the predictions on the original scale before the computation of RMSE. This provides an unbiased approach for comparing predictive performance of different models. All statistical analyses were performed using R version 4.2.3 (R Core Team 2022).

### Use of SARS-CoV-2 Concentrations

Denote the COVID-19 case count at time *t* by *C*_*t*_ and denote the SARS-CoV-2 concentrations (either O or N) measured in a wastewater sample at time *t* by *W*_*t*_. In this section, we compare different linear regression models for predicting *C*_*t*_ from *W*_*t*_. As mentioned in section 1, virus concentrations are commonly normalized by either flow, 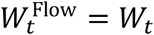 · Flow_*t*_ or a fecal marker such as PMMoV: 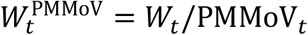. However, there have been contradictory findings on whether normalization of virus concentration can improve correlations with cases (Duvallet et al. 2022; Feng et al. 2021; Maal-Bared et al. 2023). Moreover, it seems that no studies examined using both flow and PMMoV to normalize the virus concentration, as in 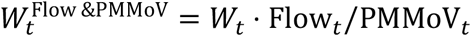. (This latter normalization has perhaps less logical coherence but to the extent that flow and PMMoV are both informative, but noisy and imperfect, it could be helpful.) We compare these three normalization approaches. It is also of interest to know whether adding lagged virus concentrations (e.g., *W*_*t*−1_ and *W*_*t*−2_) in the model will improve the predictive performance. Finally, it is prevalent in the wastewater literature to use a log_10_ transformation on the variables to meet assumptions for parametric analysis (Farkas et al. 2022; Feng et al. 2021); we compared taking the log transformation of wastewater measurements to not taking it, to validate this standard procedure. We focus on using solely the O gene here as it correlates closely with the N gene; and the S gene, while helpful in monitoring viral variants, can show gain or loss of signal accordingly. To summarize, we varied the following factors: 1. Normalization of the virus concentrations (unnormalized, flow, PMMoV or both), 2. The number of lagged values for the virus concentrations (0, 1 or 2), 3. Whether a log_10_ transformation was used on the dependent and independent variables. The three factors were ‘fully crossed’, resulting in a total of 4 ⋅ 3 ⋅ 2 *=* 24 conditions. The models were fit separately for each WWTP in each condition (we will show, shortly, that different model fits are needed for different WWTPs). To allow for comparisons between WWTPs with different sizes of population served, we divided the case count and virus concentrations by the population size. This ensures that the prediction errors for all WWTPs are theoretically on the same scale. Observations with missing values due to the creation of lagged variables were removed. As was mentioned above, the models were compared by the averaged-over-WWTPs LOOCV.

Table 3.1 displays the RMSE (root mean squared prediction error) of the cross-validated RMSE of the linear regression models under different conditions across WWTPs. Lower indicates more accurate prediction. For ease of presentation, the RMSE were multiplied by 1000. (Recall that the case count variable was divided by the corresponding catchment area population size.)

**Table 3.1:**
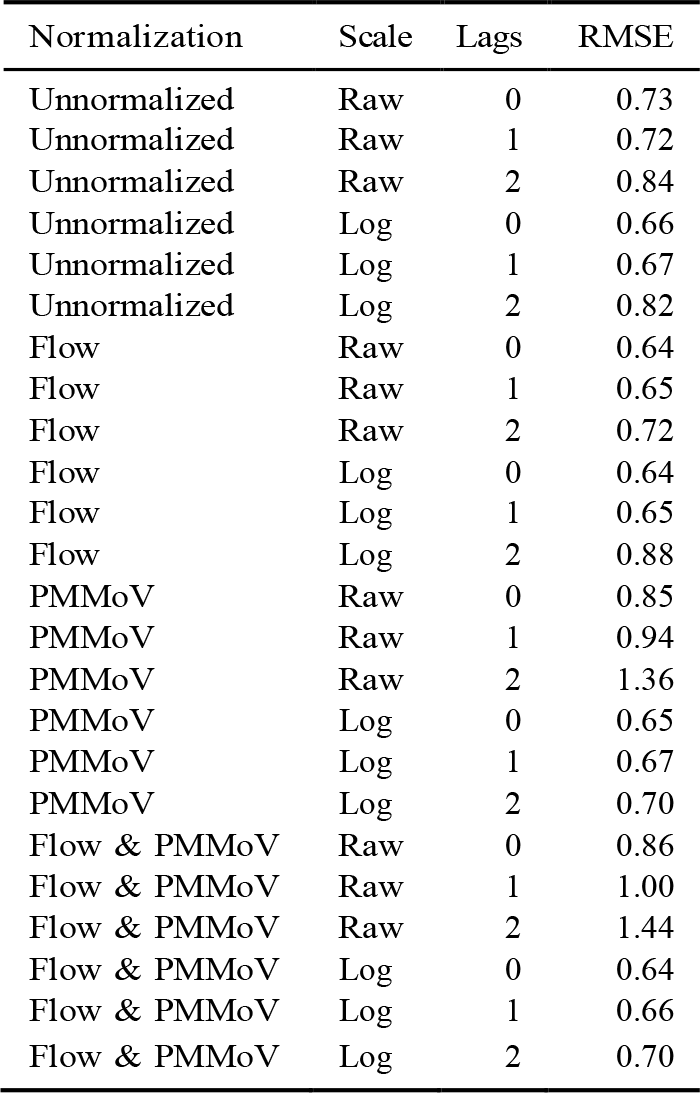
RMSEs (x1000) of RMSE across WWTPs in all 24 conditions.

| Normalization | Scale | Lags | RMSE |
| --- | --- | --- | --- |
| Unnormalized | Raw | 0 | 0.73 |
| Unnormalized | Raw | 1 | 0.72 |
| Unnormalized | Raw | 2 | 0.84 |
| Unnormalized | Log | 0 | 0.66 |
| Unnormalized | Log | 1 | 0.67 |
| Unnormalized | Log | 2 | 0.82 |
| Flow | Raw | 0 | 0.64 |
| Flow | Raw | 1 | 0.65 |
| Flow | Raw | 2 | 0.72 |
| Flow | Log | 0 | 0.64 |
| Flow | Log | 1 | 0.65 |
| Flow | Log | 2 | 0.88 |
| PMMoV | Raw | 0 | 0.85 |
| PMMoV | Raw | 1 | 0.94 |
| PMMoV | Raw | 2 | 1.36 |
| PMMoV | Log | 0 | 0.65 |
| PMMoV | Log | 1 | 0.67 |
| PMMoV | Log | 2 | 0.70 |
| Flow & PMMoV | Raw | 0 | 0.86 |
| Flow & PMMoV | Raw | 1 | 1.00 |
| Flow & PMMoV | Raw | 2 | 1.44 |
| Flow & PMMoV | Log | 0 | 0.64 |
| Flow & PMMoV | Log | 1 | 0.66 |
| Flow & PMMoV | Log | 2 | 0.70 |

Overall the results in Table 3.1 suggest that taking logs is highly important for performance; this validates the standard practice of taking log transformations not just for parameter estimation but for prediction. We find that the normalization method is not as important especially when logs are taken. When the raw data were used, normalizing by flow alone slightly reduced the RMSE, whereas involving PMMoV in the normalization led to an increase in RMSE. Recall that PMMoV is defined by truncating the highest and lowest quantile values to mitigate the high variability in its measurement; but this was not enough to remove the high level of variability totally. Our interpretation, focusing especially on the non-logged models, is that PMMoV introduces extra variability which causes predictive degradation. For instance, in our data when we normalize by PMMoV, the spike in wastewater RNA levels from the omicron surge that occurred in December 2021 in Minnesota does not appear, because the variability of the PMMoV measurements obfuscates it. Although in some contexts and for some WWTPs it seems that using PMMoV is helpful, our interpretation is that its high level of variability prevent it from being used uniformly in normalization. On the other hand, our findings validate the use of flow in that even without a log transformation, normalization by flow is seen to achieve nearly optimal predictive performance.

Including extra lags generally worsened the predictive performance, perhaps surprisingly. These results are in the context of only having a few dozen observations per WWTP, but we argue this is common given that over a longer time horizon the parameters and prediction models are likely to change in the face of evolving virus and immunity landscape. It is important to note that different models yielded different best fits for individual WWTPs; here we are presenting results averaged over all WWTPs. Overall results are affected both by higher variance and lower variance WWTPs. It is worth noting that in some of the lower variance WWTPs, predictive performance was improved by using extra lags. For instance, in the EB, EP, HT, MH, MK, and TC WWTPs, including lagged variables improves performance according to LOOCV.

Overall, the model with the lowest RMSE of the RMSEs is the one using the virus concentrations of O, normalized by flow, with log_10_ transformation, and without any lags:

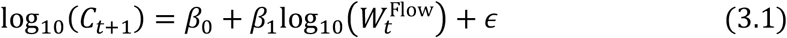

where *β*_*j*_ ‘s are the regression coefficients, and *ϵ* is the residual. Although it is not clear from our results if the normalization by flow is important, this is the model to which we explore various extensions in the upcoming sections.

#### 3.2.1 Heterogeneity in fit and individual WWTP validation

It would be ideal if it were possible to develop one model using the pooled data from all WWTPs. Unfortunately our results suggest this is not possible; we compared the estimated error variance and the estimated regression coefficients of 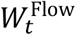 from the model (3.1) across WWTPs. We found significantly different regression coefficients and variance estimates, see Tables 3.2 (giving estimated regression coefficients and *p*-values of 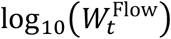 for each WWTP) and 3.3 (giving estimated *σ*^2^ values for each WWTP). We found that the estimated coefficients ranged from -0.06 to 0.66 (with summary statistics of mean = 0.28, SD = 0.23). The ratio of the largest to smallest variance estimate was larger than 10. This indicates that separate fits are needed for separate WWTPs and more broadly it suggests the important finding that in any large-scale surveillance plan, measurements from different WWTPs (even when analyzed by the same lab) may not be directly comparable and may require different interpretation.

**Table 3.2:** Estimated Coefficients and *p*-values of log_10_(*W* ^flow^) for each WWTP.

| Code | Estimated Coefficient | $p$-value | Code | Estimated Coefficient | $p$-value |
| --- | --- | --- | --- | --- | --- |
| AL | 0.47 | 0.00 | LF | 0.14 | 0.18 |
| AX | 0.13 | 0.06 | LS | 0.18 | 0.15 |
| BI | 0.10 | 0.36 | LY | -0.06 | 0.21 |
| BL | 0.48 | 0.00 | MH | 0.13 | 0.02 |
| BP | 0.20 | 0.38 | MK | 0.66 | 0.00 |
| CB | -0.01 | 0.90 | ML | 0.08 | 0.56 |
| CM | 0.09 | 0.48 | MR | 0.08 | 0.11 |
| CR | 0.36 | 0.07 | NB | 0.31 | 0.00 |
| EB | 0.21 | 0.03 | NF | 0.54 | 0.00 |
| EM | 0.59 | 0.00 | NP | 0.35 | 0.01 |
| EP | 0.61 | 0.00 | RC | 0.65 | 0.00 |
| ER | -0.02 | 0.72 | SD | 0.34 | 0.00 |
| FF | 0.06 | 0.50 | SN | 0.58 | 0.00 |
| GC | 0.03 | 0.65 | SV | 0.59 | 0.01 |
| GL | 0.19 | 0.05 | TC | 0.63 | 0.00 |
| HS | 0.28 | 0.04 | TR | 0.19 | 0.05 |
| HT | 0.09 | 0.56 | WA | 0.35 | 0.00 |
| HY | 0.16 | 0.17 | WL | -0.06 | 0.73 |
| IF | 0.61 | 0.00 | WM | 0.41 | 0.01 |
| LB | -0.03 | 0.71 | WT | 0.63 | 0.00 |

**Table 3.3:**
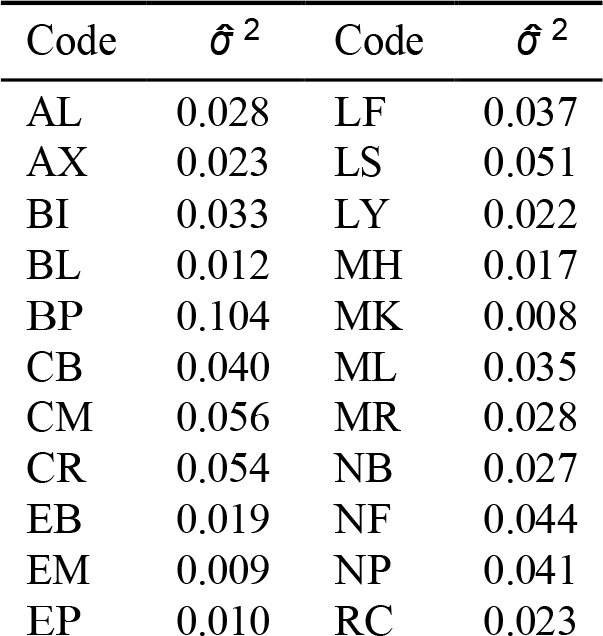
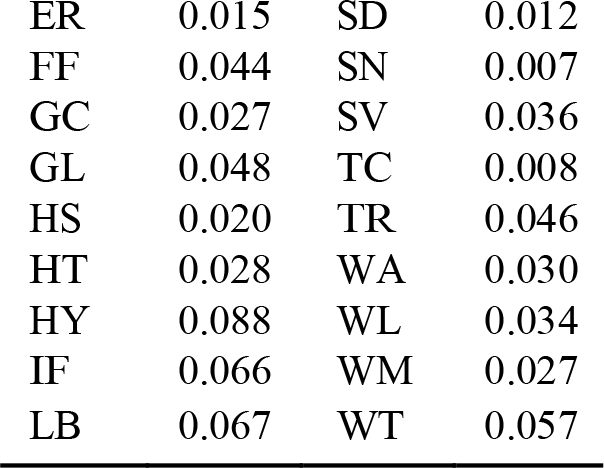
Sigma squared estimates for each WWTP.

#### 3.2.2 Use of Vaccination Rate

We examine how best to incorporate vaccination data in our linear regression models. Let 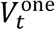 be the percentage of people who have received at least one doses of vaccine out of the population served by a WWTP, and 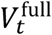 be the percentage of fully vaccinated people. The models to be investigated were built upon the model in Equation (3.1). In addition to the normalized virus concentration 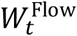, the models also considered the main effects of 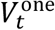 and/or 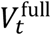, as well as two- or three-way interactions between 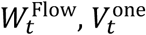 and 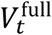. The models are listed in Table 3.4, along with their means and standard deviations of the cross-validated RMSE across WWTPs. The model that includes the interaction between 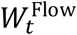 and 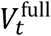,

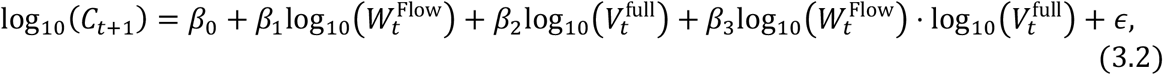

minimizes the mean RMSE, but the differences are not large and the results were not sensitive to whether 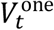 or 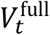 was used. The clearest result is that including too many variables leads to worse predictions. However, as detailed in the next subsection we observe that including vaccination may add more variability than it removes (especially when predicting two weeks ahead).

**Table 3.4:** Means and standard deviations (x1000) of RMSE across WWTPs for different vaccination linear models.

| Formula | Mean | SD |
| --- | --- | --- |
| $\log_{10}(C_{t+1}) \sim \log_{10}(W^{\text{flow}}) + \log_{10}(V^{\text{one}})$ | 0.55 | 0.21 |
| $\log_{10}(C_{t+1}) \sim \log_{10}(W^{\text{flow}}) + \log_{10}(V^{\text{full}})$ | 0.54 | 0.23 |
| $\log_{10}(C_{t+1}) \sim \log_{10}(W^{\text{flow}}) + \log_{10}(V^{\text{one}}) + \log_{10}(V^{\text{full}})$ | 2.47 | 12.26 |
| $\log_{10}(C_{t+1}) \sim \log_{10}(W^{\text{flow}}) * \log_{10}(V^{\text{one}})$ | 0.54 | 0.20 |
| $\log_{10}(C_{t+1}) \sim \log_{10}(W^{\text{flow}}) * \log_{10}(V^{\text{full}})$ | 0.53 | 0.22 |
| $\log_{10}(C_{t+1}) \sim \log_{10}(W^{\text{flow}}) * \log_{10}(V^{\text{one}}) * \log_{10}(V^{\text{full}})$ | 149.12 | 938.86 |

#### 3.2.3 Prediction Accuracy Over Different Forecast Horizons

We assess the performance of predictions over different forecast horizons (same week, one week ahead and two weeks ahead). The model in Equation (3.2) was fitted separately for each WWTP, with log_10_(*C*_*t*_), log_10_(*C*_*t*+1_) and log_10_(*C*_*t*+2_) being the dependent variable. The means and standard deviations of the cross-validated RMSE are reported in the first three rows of Table 3.5. At the current noise level, we do not see a degradation in ability to predict one week ahead versus predicting the current week’s case count. The predictions for two weeks ahead is substantially worse than the predictions for the first week. For comparisons, we fitted the model in Equation (3.1) again with different forecast horizons (last three rows of Table 3.5). Interestingly, when predicting two weeks ahead, the simplest model (excluding vaccination information) was more accurate in terms of both mean and variance.

**Table 3.5:** Means and SDs (x1000) of RMSE across WWTPs when predicting the case count of the same week, one week ahead and two weeks ahead.

| Model | Mean | SD |
| --- | --- | --- |
| $\log(C_t) \sim \log_{10}(W_t^{\text{flow}}) * \log_{10}(V_t^{\text{full}})$ | 0.54 | 0.21 |
| $\log(C_{t+1}) \sim \log_{10}(W_t^{\text{flow}}) * \log_{10}(V_t^{\text{full}})$ | 0.53 | 0.22 |
| $\log(C_{t+2}) \sim \log_{10}(W_t^{\text{flow}}) * \log_{10}(V_t^{\text{full}})$ | 0.75 | 1.47 |
| $\log(C_t) \sim \log_{10}(W_t^{\text{flow}})$ | 0.59 | 0.19 |
| $\log(C_{t+1}) \sim \log_{10}(W_t^{\text{flow}})$ | 0.61 | 0.19 |
| $\log(C_{t+2}) \sim \log_{10}(W_t^{\text{flow}})$ | 0.60 | 0.19 |

## 4 Storage and Time-to-Analysis Stability Study Findings

Although one might expect a priori for measurements from different WWTPs that that are studied in the same lab to have broadly similar characteristics, we found above, in the analysis of our surveillance data, nontrivial heterogeneity in predictive ability (coefficient values and standard deviation estimates) between different WWTPs. Unmeasured variables such as population mobility, access to clinical testing centers, or population willingness to utilize clinical testing could account for some of the variation. Wastewater storage conditions are another possible cause. To shed light on what causes such unexplained large differences between WWTPs, we conducted a stability study using a large volume of WW from a single site that was partitioned into samples that were stored at varying conditions (4C, -20, and -80C) for 2,5 7, or 14 days. Nucleic acid for each sample (n=4) was then analyzed by qRT PCR concurrently (for a total of 4 * 4 * 5 = 80 data points, with summary statistic means (SDs) for each temperature [averaging across all times] of 39400 (8560) for baseline, 18600 (8600) for 4C, 2970 (5420) for -20C, and 3360 (1550) for -80C)

We found that storage temperature had a large and significant impact on the measured RNA. The length of time of storage had a less noticeable and not significant impact. We subtracted each baseline measurement from each storage measurement, yielding a negative number in all cases. We transformed the response by taking the negative and then the logarithm. We then fit a linear mixed regression model with no intercept term, a (linear) time trend term, a factor variable for the three storage conditions, and a random intercept effect for each sampling unit, with no interactions included; that is, we fit

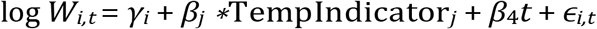

for *i* = 1, …, 4, *t ∈ {*2, 5, 7, 14*}*, TempIndicator_*j*_ an indicator/dummy variable for the three temperature storage conditions, *γ*_*i*_ i.i.d. *N* (0, *σ*^2^) random effects, *σ*^2^ *>* 0, *ϵ*_*i,t*_ i.i.d. *N* (0, *η*^2^), and *β*_*j*_ the fixed effects of interest. The coefficient estimates for each of the three storage conditions were significant; the coefficients for −20*C* and −80*C* were not significantly different than each other, but are different than the coefficient for 4*C*. The negative exponentiated coefficient estimates (p-values) are -16200 (2 *×* 10^−12^) for 4C, -32300 (1 *×* 10^−14^) for -20C, and -32300 (1 *×* 10^−14^) for -80C. The time trend term was not significant. The means and standard deviations (over the four samples) at each of the 12 different conditions are presented in Table 4.1. The Day 0 average (sd) is 39400 (8560). It is easy to see at a glance the effect of storing below 4 C.

**Table 4.1:** Means (standard deviations) of RNA levels over 4 samples for each of 4 storage lengths at 3 different temperatures.

|  | Day 2 | Day 5 | Day 7 | Day 14 |
| --- | --- | --- | --- | --- |
| 4 C | 21900 (6100) | 22000 (11700) | 15900 (11200) | 14500 (2700) |
| -20 C | 900 (200) | 4700 (7600) | 5600 (7900) | 700 (500) |
| -80 C | 4000 (1100) | 2800 (1300) | 3600 (2300) | 3000 (1600) |

These data provide important guidelines for sample submission and storage whereby 4C stored samples showed less degradation than a -20C or -80C freeze and thaw. Under these principles we advocate long term storage at -80C and shipping temperatures under refrigeration (4C).

## 5 Conclusions

We studied surveillance data for SARS-CoV-2 across the state of Minnesota and undertook efforts to apply statistical analyses to inform how to best interpret and apply wastewater data by studying the predictive relationship of wastewater virus levels on clinical case counts. We acknowledge that using clinical case counts as a validation benchmark is an imperfect solution, namely because wastewater is in fact intended to solve some of the problems that arise from using clinical case counts, as was discussed above. Nonetheless, since case counts are the baseline standard used for public health decision making, they serve as a natural benchmark. For instance, it is clear that if clinical case counts demonstrate a large surge, then the ability of wastewater to demonstrate the same surge is important and beneficial; so in this sense a strong statistical relationship between the wastewater measurements and case counts is meaningful.

Our estimates of error variance and regression coefficients vary substantially across WWTPs. These findings suggest that the relationship between COVID-19 incidence and SARS-CoV-2 RNA in wastewater may be treatment plant-specific, and future work will need to continue investigating how to appropriately normalize data from different plants to allow for cross-plant comparisons. Additionally, this suggests that at present, COVID-19 WBE may need to be viewed at individual plant level for its most impactful application. The variation is not accounted for by the catchment area or the size of the corresponding population.

Prompted by this variability we set out to dissect this phenomena to determine if we could identify actionable items that would boost accuracy (ie diminish variance). Toward that, we show that wastewater RNA are quite sensitive to storage conditions, so such storage conditions are a strong candidate for accounting for at least some of the variation.

Consistent with some studies in the wastewater literature (Maal-Bared et al. 2023; Feng et al. 2021; Duvallet et al. 2022), the current study finds that the predictive performance does not improve after involving PMMoV in the normalization. Interestingly, adding lagged virus concentrations to the model generally worsened the predictive performance. Using the vaccination rate in the model has been shown to improve the predictions. However, the results are not very consistent across different forecast horizons. Due to the relatively low number of data points for each WWTP (recall Table 2.1), nonlinear or machine learning methods are not effective, and in fact often very parsimonious linear regression models performed best. One shortcoming of this study is that the available data are rather limited, and the conclusions may not apply in an extremely data rich environment. However, in infectious disease modeling, we believe that true data rich environments (in which necessarily model parameters and immunological landscapes all stay constant over time) are exceedingly rare. As such, based on data from a broad geographic area studied during the SARS-CoV2 pandemic, we detail our findings that are additive to the fields of epidemiology and statistical modeling and prediction, and which provide new guidance for storage procedures for wastewater samples.

## Data Availability

All data produced in the present study are available upon reasonable request to the authors

## Acknowledgements

This work was funded, in part, by a grant from 3M and the Minnesota Department of Health.

## Notes

### Competing Interest Statement

The authors have declared no competing interest.

## Reference

Ahmed, Warish, Nicola Angel, Janette Edson, Kyle Bibby, Aaron Bivins, Jake W O’Brien, Phil M Choi, et al. 2020. “First Confirmed Detection of SARS-CoV-2 in Untreated Wastewater in Australia: A Proof of Concept for the Wastewater Surveillance of COVID-19 in the Community.” Science of the Total Environment 728: 138764.

Arora, Sudipti, Aditi Nag, Jasmine Sethi, Jayana Rajvanshi, Sonika Saxena, Sandeep K Shrivastava, and Akhilendra Bhushan Gupta. 2020. “Sewage Surveillance for the Presence of SARS-CoV-2 Genome as a Useful Wastewater Based Epidemiology (WBE) Tracking Tool in India.” Water Science and Technology 82 (12): 2823–36.

Barasa, Edwine W, Paul O Ouma, and Emelda A Okiro. 2020. “Assessing the Hospital Surge Capacity of the Kenyan Health System in the Face of the COVID-19 Pandemic.” PLoS One 15 (7): e0236308.

Chan, Vinson Wai-Shun, Peter Ka-Fung Chiu, Chi-Hang Yee, Yuhong Yuan, Chi-Fai Ng, and Jeremy Yuen-Chun Teoh. 2021. “A Systematic Review on COVID-19: Urological Manifestations, Viral RNA Detection and Special Considerations in Urological Conditions.” World Journal of Urology 39 (9): 3127–38.

Chen, Yifei, Liangjun Chen, Qiaoling Deng, Guqin Zhang, Kaisong Wu, Lan Ni, Yibin Yang, et al. 2020. “The Presence of SARS-CoV-2 RNA in the Feces of COVID-19 Patients.” Journal of Medical Virology 92 (7): 833–40.

Cheung, Ka Shing, Ivan FN Hung, Pierre PY Chan, KC Lung, Eugene Tso, Raymond Liu, YY Ng, et al. 2020. “Gastrointestinal Manifestations of SARS-CoV-2 Infection and Virus Load in Fecal Samples from a Hong Kong Cohort: Systematic Review and Meta-Analysis.” Gastroenterology 159 (1): 81–95.

Duvallet, Claire, Fuqing Wu, Kyle A McElroy, Maxim Imakaev, Noriko Endo, Amy Xiao, Jianbo Zhang, et al. 2022. “Nationwide Trends in COVID-19 Cases and SARS-CoV-2 RNA Wastewater Concentrations in the United States.” ACS ES&T Water.

ESRI 2023. ArcGIS Online, Esri Demographics. Redlands, CA: Environmental Systems Research Institute.

Farkas, Kata, Cameron Pellett, Natasha Alex-Sanders, Matthew TP Bridgman, Alexander Corbishley, Jasmine MS Grimsley, Barbara Kasprzyk-Hordern, et al. 2022. “Comparative Assessment of Filtration-and Precipitation-Based Methods for the Concentration of SARS-CoV-2 and Other Viruses from Wastewater.” Microbiology Spectrum 10 (4): e01102–22.

Feng, Shuchen, Adelaide Roguet, Jill S McClary-Gutierrez, Ryan J Newton, Nathan Kloczko, Jonathan G Meiman, and Sandra L McLellan. 2021. “Evaluation of Sampling, Analysis, and Normalization Methods for SARS-CoV-2 Concentrations in Wastewater to Assess COVID-19 Burdens in Wisconsin Communities.” Acs Es&T Water 1 (8): 1955–65.

Hamza, Hazem, Neveen Magdy Rizk, Mahmoud Afw Gad, and Ibrahim Ahmed Hamza. 2019. “Pepper Mild Mottle Virus in Wastewater in Egypt: A Potential Indicator of Wastewater Pollution and the Efficiency of the Treatment Process.” Archives of Virology 164 (11): 2707–13.

Hasan, Shadi W, Yazan Ibrahim, Marianne Daou, Hussein Kannout, Nila Jan, Alvaro Lopes, Habiba Alsafar, and Ahmed F Yousef. 2021. “Detection and Quantification of SARS-CoV-2 RNA in Wastewater and Treated Effluents: Surveillance of COVID-19 Epidemic in the United Arab Emirates.” Science of The Total Environment 764: 142929.

Hellmér, Maria, Nicklas Paxéus, Lars Magnius, Lucica Enache, Birgitta Arnholm, Annette Johansson, Tomas Bergström, and Heléne Norder. 2014. “Detection of Pathogenic Viruses in Sewage Provided Early Warnings of Hepatitis a Virus and Norovirus Outbreaks.” Applied and Environmental Microbiology 80 (21): 6771–81.

Khan, Kamruzzaman, Scott W Tighe, and Appala Raju Badireddy. 2021. “Factors Influencing Recovery of SARS-CoV-2 RNA in Raw Sewage and Wastewater Sludge Using Polyethylene Glycol–Based Concentration Method.” Journal of Biomolecular Techniques: JBT 32 (3): 172.

Kitajima, Masaaki, Brandon C Iker, Ian L Pepper, and Charles P Gerba. 2014. “Relative Abundance and Treatment Reduction of Viruses During Wastewater Treatment Processes—Identification of Potential Viral Indicators.” Science of the Total Environment 488: 290–96.

Kitajima, Masaaki, Hannah P Sassi, and Jason R Torrey. 2018. “Pepper Mild Mottle Virus as a Water Quality Indicator.” NPJ Clean Water 1 (1): 1–9.

Lauer, Stephen A, Kyra H Grantz, Qifang Bi, Forrest K Jones, Qulu Zheng, Hannah R Meredith, Andrew S Azman, Nicholas G Reich, and Justin Lessler. 2020. “The Incubation Period of Coronavirus Disease 2019 (COVID-19) from Publicly Reported Confirmed Cases: Estimation and Application.” Annals of Internal Medicine 172 (9): 577–82.

Li, Ruiyun, Sen Pei, Bin Chen, Yimeng Song, Tao Zhang, Wan Yang, and Jeffrey Shaman. 2020. “Substantial Undocumented Infection Facilitates the Rapid Dissemination of Novel Coronavirus (SARS-CoV-2).” Science 368 (6490): 489–93.

Maal-Bared, Rasha, Yuanyuan Qiu, Qiaozhi Li, Tiejun Gao, Steve E Hrudey, Sudha Bhavanam, Norma J Ruecker, Erik Ellehoj, Bonita E Lee, and Xiaoli Pang. 2023. “Does Normalization of SARS-CoV-2 Concentrations by Pepper Mild Mottle Virus Improve Correlations and Lead Time Between Wastewater Surveillance and Clinical Data in Alberta (Canada): Comparing Twelve SARS-CoV-2 Normalization Approaches.” Science of The Total Environment 856: 158964.

Manor, Y, R Handsher, T Halmut, M Neuman, A Bobrov, H Rudich, A Vonsover, L Shulman, O Kew, and E Mendelson. 1999. “Detection of Poliovirus Circulation by Environmental Surveillance in the Absence of Clinical Cases in Israel and the Palestinian Authority.” Journal of Clinical Microbiology 37 (6): 1670–75.

Melvin, Richard G, Emily N Hendrickson, Nabiha Chaudhry, Onimitein Georgewill, Rebecca Freese, Timothy W Schacker, and Glenn E Simmons Jr. 2021. “A Novel Wastewater-Based Epidemiology Indexing Method Predicts SARS-CoV-2 Disease Prevalence Across Treatment Facilities in Metropolitan and Regional Populations.” Scientific Reports 11 (1): 21368.

Nishiura, Hiroshi, Tetsuro Kobayashi, Takeshi Miyama, Ayako Suzuki, Sung-mok Jung, Katsuma Hayashi, Ryo Kinoshita, et al. 2020. “Estimation of the Asymptomatic Ratio of Novel Coronavirus Infections (COVID-19).” International Journal of Infectious Diseases 94: 154–55.

Oran, Daniel P, and Eric J Topol. 2020. “Prevalence of Asymptomatic SARS-CoV-2 Infection: A Narrative Review.” Annals of Internal Medicine 173 (5): 362–67.

Parasa, Sravanthi, Madhav Desai, Viveksandeep Thoguluva Chandrasekar, Harsh K Patel, Kevin F Kennedy, Thomas Roesch, Marco Spadaccini, et al. 2020. “Prevalence of Gastrointestinal Symptoms and Fecal Viral Shedding in Patients with Coronavirus Disease 2019: A Systematic Review and Meta-Analysis.” JAMA Network Open 3 (6): e2011335–35.

Peccia, Jordan, Alessandro Zulli, Doug E Brackney, Nathan D Grubaugh, Edward H Kaplan, Arnau Casanovas-Massana, Albert I Ko, et al. 2020. “Measurement of SARS-CoV-2 RNA in Wastewater Tracks Community Infection Dynamics.” Nature Biotechnology 38 (10): 1164–67.

Post, Lori Ann, Tariq Ziad Issa, Michael J Boctor, Charles B Moss, Robert L Murphy, Michael G Ison, Chad J Achenbach, et al. 2020. “Dynamic Public Health Surveillance to Track and Mitigate the US COVID-19 Epidemic: Longitudinal Trend Analysis Study.” Journal of Medical Internet Research 22 (12): e24286.

R Core Team. 2022. R: A Language and Environment for Statistical Computing. Vienna, Austria: R Foundation for Statistical Computing. https://www.R-project.org/.

Randazzo, Walter, Pilar Truchado, Enric Cuevas-Ferrando, Pedro Simón, Ana Allende, and Gloria Sánchez. 2020. “SARS-CoV-2 RNA in Wastewater Anticipated COVID-19 Occurrence in a Low Prevalence Area.” Water Research 181: 115942.

Rosario, Karyna, Erin M Symonds, Christopher Sinigalliano, Jill Stewart, and Mya Breitbart. 2009. “Pepper Mild Mottle Virus as an Indicator of Fecal Pollution.” Applied and Environmental Microbiology 75 (22): 7261–67.

Weidhaas, Jennifer, Zachary T Aanderud, D Keith Roper, James VanDerslice, Erica Brown Gaddis, Jeff Ostermiller, Ken Hoffman, et al. 2021. “Correlation of SARS-CoV-2 RNA in Wastewater with COVID-19 Disease Burden in Sewersheds.” Science of The Total Environment 775: 145790.

Wong, Martin CS, Junjie Huang, Christopher Lai, Rita Ng, Francis KL Chan, and Paul KS Chan. 2020. “Detection of SARS-CoV-2 RNA in Fecal Specimens of Patients with Confirmed COVID-19: A Meta-Analysis.” Journal of Infection 81 (2): e31–38.

Ye, Yinyin, Robert M Ellenberg, Katherine E Graham, and Krista R Wigginton. 2016. “Survivability, Partitioning, and Recovery of Enveloped Viruses in Untreated Municipal Wastewater.” Environmental Science & Technology 50 (10): 5077–85.

Zhan, Qingyu, Kristina M Babler, Mark E Sharkey, Ayaaz Amirali, Cynthia C Beaver, Melinda M Boone, Samuel Comerford, et al. 2022. “Relationships Between SARS-CoV-2 in Wastewater and COVID-19 Clinical Cases and Hospitalizations, with and Without Normalization Against Indicators of Human Waste.” ACS ES&T Water.

Zhang, Tao, Mya Breitbart, Wah Heng Lee, Jin-Quan Run, Chia Lin Wei, Shirlena Wee Ling Soh, Martin L Hibberd, Edison T Liu, Forest Rohwer, and Yijun Ruan. 2006. “RNA Viral Community in Human Feces: Prevalence of Plant Pathogenic Viruses.” PLoS Biology 4 (1): e3.

